# Whole genome sequence analysis of *Salmonella* Typhi in Papua New Guinea reveals an established population of genotype 2.1.7 sensitive to antimicrobials

**DOI:** 10.1101/2021.09.27.21264209

**Authors:** Zoe A. Dyson, Elisheba Malau, Paul F. Horwood, Rebecca Ford, Valentine Siba, Mition Yoannes, William Pomat, Megan Passey, Louise M. Judd, Danielle J. Ingle, Deborah A. Williamson, Gordon Dougan, Andrew R. Greenhill, Kathryn E. Holt

## Abstract

**Background:** Typhoid fever, a systemic infection caused by *Salmonella enterica* serovar Typhi, remains a considerable public health threat in impoverished regions within many low- and middle-income settings. However, we still lack a detailed understanding of the emergence, population structure, molecular mechanisms of antimicrobial resistance (AMR), and transmission dynamics of *S*. Typhi across many settings, particularly throughout the Asia-Pacific islands. Here we present a comprehensive whole genome sequence (WGS) based overview of *S*. Typhi populations circulating in Papua New Guinea (PNG) over 30 years.

**Principle findings:** Bioinformatic analysis of 86 *S*. Typhi isolates collected between 1980-2010 demonstrated that the population structure of PNG is dominated by a single genotype (2.1.7) that appears to have emerged in the Indonesian archipelago in the mid-twentieth century with very limited evidence of inter-country transmission. Genotypic and phenotypic data demonstrated that the PNG *S*. Typhi population appears to be susceptible to former first line drugs for treating typhoid fever (chloramphenicol, ampicillin and co-trimoxazole), as well as fluoroquinolones, third generation cephalosporins, and macrolides. PNG genotype 2.1.7 was genetically conserved, with very few deletions, and no evidence of plasmid or prophage acquisition. Genetic variation among this population was attributed to either single point mutations, or homologous recombination adjacent to repetitive ribosomal RNA operons.

**Significance:** Antimicrobials remain an effective option for the treatment of typhoid fever in PNG, along with other intervention strategies including improvements to water, sanitation and hygiene (WaSH) related infrastructure and potentially the introduction of Vi-conjugate vaccines. However, continued genomic surveillance is warranted to monitor for the emergence of AMR within local populations, or the introduction of AMR associated genotypes of *S*. Typhi in this setting.

**Author Summary:** Typhoid fever, caused by *Salmonella enterica* serovar Typhi, is a systemic infection common to many low- to middle-income settings. While the population structure of *S*. Typhi has been genetically characterised using whole genome sequencing in many endemic countries throughout Sub-Saharan Africa and South Asia, we are still lacking a detailed understanding for many regions including those among the Asia-Pacific islands. Genomic surveillance of isolates spanning 30 years demonstrated a population structure of *S*. Typhi in Papua New Guinea (PNG) dominated by a single genotype (2.1.7) that emerged in the mid-twentieth century, is genetically homogeneous, and sensitive to a wide range of antibiotics commonly used in the treatment of typhoid. There was little evidence of inter-country transmission and the setting appeared free of *S*. Typhi genotypes commonly associated with AMR e.g. H58 (genotype 4.3.1). These data suggest that former first line drugs (chloramphenicol, ampicillin and co-trimoxazole), fluoroquinolones, third generation cephalosporins and macrolides all remain viable options for controlling typhoid in addition to the introduction of Vi-conjugate vaccines and improvements to water, sanitation and hygiene (WaSH) related infrastructure. Routine molecular surveillance is necessary to monitor for introduced or emerging AMR to inform treatment guidelines and intervention strategies.

## Introduction

Typhoid fever is a systemic infection of the bacterium *Salmonella enterica* serovar Typhi[1]. Each year more than 10,000,000 cases of typhoid fever occur worldwide, of which more than 100,000 result in death[2]. *S*. Typhi is transmitted faeco-orally, usually through contaminated food and water and therefore constitutes a major public health threat in many low- to middle-income countries (LMIC), particularly in impoverished regions where hygiene and sanitation infrastructure is limited[3].

Antimicrobial chemotherapy has become a mainstay in the treatment of typhoid fever, with death rates declining from 20-30% to 1% following the introduction of antimicrobials[4,5]. However, antimicrobial resistance (AMR) has arisen multiple times in *S*. Typhi, with multidrug-resistance (MDR; resistance to former first line antimicrobials chloramphenicol, co-trimoxazole, and ampicillin) emerging from the 1970s onwards[6], followed by resistance to fluoroquinolones commonly mediated by mutations in the quinolone resistance determining region (QRDR) of genes *gyrA, gyrB*, and *parC*[7–9]. More recently, extensively drug-resistant (XDR) *S*. Typhi populations resistant to all orally administered antimicrobials except for azithromycin have emerged in Pakistan outbreaks[10,11], and mutations mediating azithromycin resistance have been observed in non-XDR *S*. Typhi populations[12–15].

Papua New Guinea (PNG) has been recognised as a high burden setting for typhoid since the mid-1990s when the incidence rate was reported as 1,208 cases per 100,000 population, amongst the highest rates in the world at that time[16]. Although there is a paucity of recent data, one diagnostic study demonstrated that typhoid remains a common diagnosis in febrile patients in the PNG highlands[17]. Vaccines for typhoid fever are not currently part of the routine schedule in PNG, nor have they been in the past.

Early molecular studies [18–20] revealed that several ribotypes of *S*. Typhi appeared endemic to PNG, with many appearing unique to the country, and some also observed in Southeast Asian countries. Further, 80.6% of isolates from 1986-1989 belonged to a single phage type (D2) [18,20], which was also observed at lower frequency in other countries including Malaysia and Thailand. Previous reports[18], have also noted that *S*. Typhi from PNG appeared free of both detectable AMR genes and plasmids, and have suggested that genome plasticity might be common. However, despite these early observations, only two locally collected *S*. Typhi sequences from PNG have been reported on in detail[21]. A further 45 isolates collected from travellers returning to Australia from PNG were sequenced as a part of a global overview study[9], but this study did not report on the specific dynamics and characteristics of the PNG pathogen population. Subsequently, there is currently no comprehensive whole genome sequence-based overview of the population structure, transmission dynamics, or evolutionary history of *S*. Typhi in PNG. Here we combine data on 41 novel isolates collected locally in PNG between 1992 and 2010, with those data sequenced previously from return travellers, to provide a 30-year overview (1980-2010) of *S*. Typhi in PNG.

## Methods

### Ethics statement

Ethical approval for this study was granted by the PNG Institute of Medical Research Institutional Review Board (1609), the PNG Medical Research Advisory Board (MRAC 16.43) and the Federation University Human Research Ethics Committee (A17-074).

### Study Setting

PNG is a tropical country situated approximately 6° south of the equator in the Western Pacific region. It is a LMIC, with a large proportion of its ∼9 million inhabitants living in regional and remote areas of the country. Health and development indicators are low by regional and global standards, with ∼40% of people having access to improved water supply and <20% with access to adequate sanitation[22,23]. In 2019, PNG’s human development index of 0.55 ranked it 155 out of 189 countries and territories[24].

The PNG Institute of Medical Research (PNGIMR) is headquartered in Goroka, the capital of Eastern Highlands Province. The institute has been responsible for the majority of studies that have been conducted on typhoid fever in PNG.

### Isolate collection and culture processing

Isolates of *S*. Typhi that were sequenced as part of this study (n=41) were obtained from the PNGIMR culture collection. All isolates had been preserved by freeze-drying in glass ampules. The content of each ampule was resuspended in nutrient broth, and inoculated onto nutrient agar (incubated for 18 – 24 hours at 35°C). PCR was conducted on isolates using previously described methods [17] to confirm their identity as *S*. Typhi.

### Antimicrobial susceptibility testing and phage typing

Antimicrobial susceptibility testing (AST) was carried out for n=40 (88.9%) of travel-associated cases for chloramphenicol, ampicillin, sulfamethoxazole, trimethoprim, nalidixic acid, tetracycline, kanamycin, gentamycin, and spectinomycin as detailed in **S1 Table**. AST was performed on *S*. Typhi isolates using agar breakpoint dilution and interpreted using Clinical and Laboratory Standards Institute (CLSI) breakpoints as described previously[25]. Phage types were extracted from the National Enteric Pathogens Surveillance Scheme (NEPSS), a dedicated surveillance system for human and nonhuman enteric pathogens (including *Salmonella*) that has been operated by the Microbiological Diagnostic Unit Public Health Laboratory (MDU PHL)[25].

### DNA extraction and whole genome sequencing

Isolated colonies (1 – 3) were suspended in sterile water to 0.5 McFarland, and 100 µl of the suspension was used for DNA extraction with the DNeasy Blood and Tissue Kit (Qiagen, Hilden, Germany), following the manufacturer’s instructions. Extracted genomic DNA (gDNA) was then subjected to indexed whole genome sequencing on an Illumina HiSeq 2500 platform at the Wellcome Sanger Institute to generate paired end reads of 150 bp in length as described previously[9].

Three isolates were selected for additional long-read sequencing using the Nanopore MinION R9 device which was carried out as described previously[26]. To generate high molecular weight gDNA (>60□kbp) suited to Oxford Nanopore sequencing without further size selection the following protocol was used; firstly, isolates were grown overnight at 37°C on Luria-Bertani (LB) plates, before single colonies were picked for overnight culture at 37°C in LB broth medium. Bacterial cell pellets from 3.0□ml LB culture were then generated by centrifugation at 15,000□g for 5□minutes. gDNA was extracted from these pellets using Agencourt GenFind V2 (Beckman Coulter) with modifications as follows. Cell pellets were resuspended in 400□µl lysis buffer containing 9□µl Proteinase K (96 mg/ml; Beckman Coulter) and 1□µl RNase A (100□mg/ml; Sigma Aldrich) by tip mixing. Samples were lysed at 37°C for 30□minutes, and gDNA was extracted from the lysed samples by completing the remaining steps of the GenFind V2 for 200□µl of blood/serum from the binding step onwards as per the manufacturer’s instructions.

### Read mapping, Single Nucleotide Variant (SNV) and phylogenomic analyses

For Single Nucleotide Variant (SNV) analysis, paired end reads for n=41 *S*. Typhi isolates collected in this study together with n=45 from a previous study[27] (**S1 Table**), were aligned to the CT18 reference genome (GenBank accession AL513382) [28] using the RedDog mapping pipeline (V1.beta10.3; available at https://github.com/katholt/reddog). RedDog maps reads to the reference genome with Bowtie2 (v2.2.9)[29], before using SAMtools (v1.3.1) [30] to identify high quality single nucleotide variant (SNV) calls as previously described[8]. A single sequence (sample MDUST355) was found to be a mixture of *S*. Typhi genotypes (∼80% 2.1.7.1 and ∼20% 2.1.7.2) and was subsequently removed from further phylogenomic analyses. A core SNV alignment was generated by concatenating alleles with high-quality consensus base calls (phred scores >20), for all SNV sites meeting this criteria in >95% of genomes (representing the 95% ‘soft’ core of the *S*. Typhi genome). This alignment was filtered to exclude SNV called in previously defined [9,31,32] prophage regions and repetitive sequences (354 kb; ∼7.4% of bases) in the CT18 reference chromosome, and Paratyphi A sequence AKU_12601 (GenBank accession FM200053) alleles were also included in the alignment for outgroup rooting of the phylogenetic tree. The resultant 657 bp SNV alignment then was used to infer a whole genome pseudoalignment using the CT18 reference sequence, and any remaining recombination was removed using Gubbins (v2.3.2)[33]. The resultant 620 bp recombination-filtered alignment was used as input for RAxML (v8.2.8)[34], which was used to infer a maximum likelihood (ML) phylogeny with a generalized time-reversible model and a Gamma distribution to model site-specific rate variation (GTR + Γ substitution model; GTRGAMMA in RAxML), with 100 bootstrap pseudo-replicates used to assess branch support (**S1 Fig**).

GenoTyphi (v1.9.1; available at https://github.com/katholt/genotyphi/) was used together with RedDog Bowtie2 alignment (BAM) files to infer genotypes for all sequences analysed in this study using SNVs that were defined in the extended *S*. Typhi genotyping framework[27,35]. These data revealed that our sampling of the PNG *S*. Typhi population was dominated by a single genotype (2.1.7, n=84, 98%; **S1 Fig**), necessitating the completion of a reference genome for the PNG genotype 2.1.7 population to allow for high resolution phylogenomic analyses. Three PNG genotype 2.1.7 genomes were selected for Nanopore long read sequencing and hybrid assembly (see **S1 Table**), with the oldest of these (MDUST348 from 1985), selected as a reference sequence for lineage-focused SNV analyses and phylogenetic inference using the mapping-based approach described above. For this newly completed PNG genotype 2.1.7 reference genome (MDUST348, deposited under accession TBA, repetitive regions were identified using the ncumer command in MUMmer (v3.23) [36] and integrated prophage sequences detected using the PHAge Search Tool Enhanced Release (PHASTER; available at https://phaster.ca/)[37]. These regions (detailed in **S2 Table**) were then filtered from the inferred PNG genotype 2.1.7 alignment, with any further recombination removed with Gubbins as described above. The resultant recombination filtered alignment of PNG genotype 2.1.7 *S*. Typhi sequences (n=83) also included alleles from representative non-genotype 2.1.7 sequences as outgroups for phylogenetic tree rooting (detailed in **S3 Table**), and this final alignment (258 bp; 143 sequences) was then used to infer the final phylogenies shown in **Fig 1 and S2 Fig**. A global context phylogeny including an additional n=50 sequences of clade 2.1 (n=133 total; **S1 Table**) from previous studies[27,38] was inferred in the same manner with an alignment of 671 bp in length (**Fig 2**; 184 sequences including outgroups). An interactive version of the resulting global context clade 2.1 phylogeny is available at: https://microreact.org/project/icGPbF_O8.

**Fig 1.**
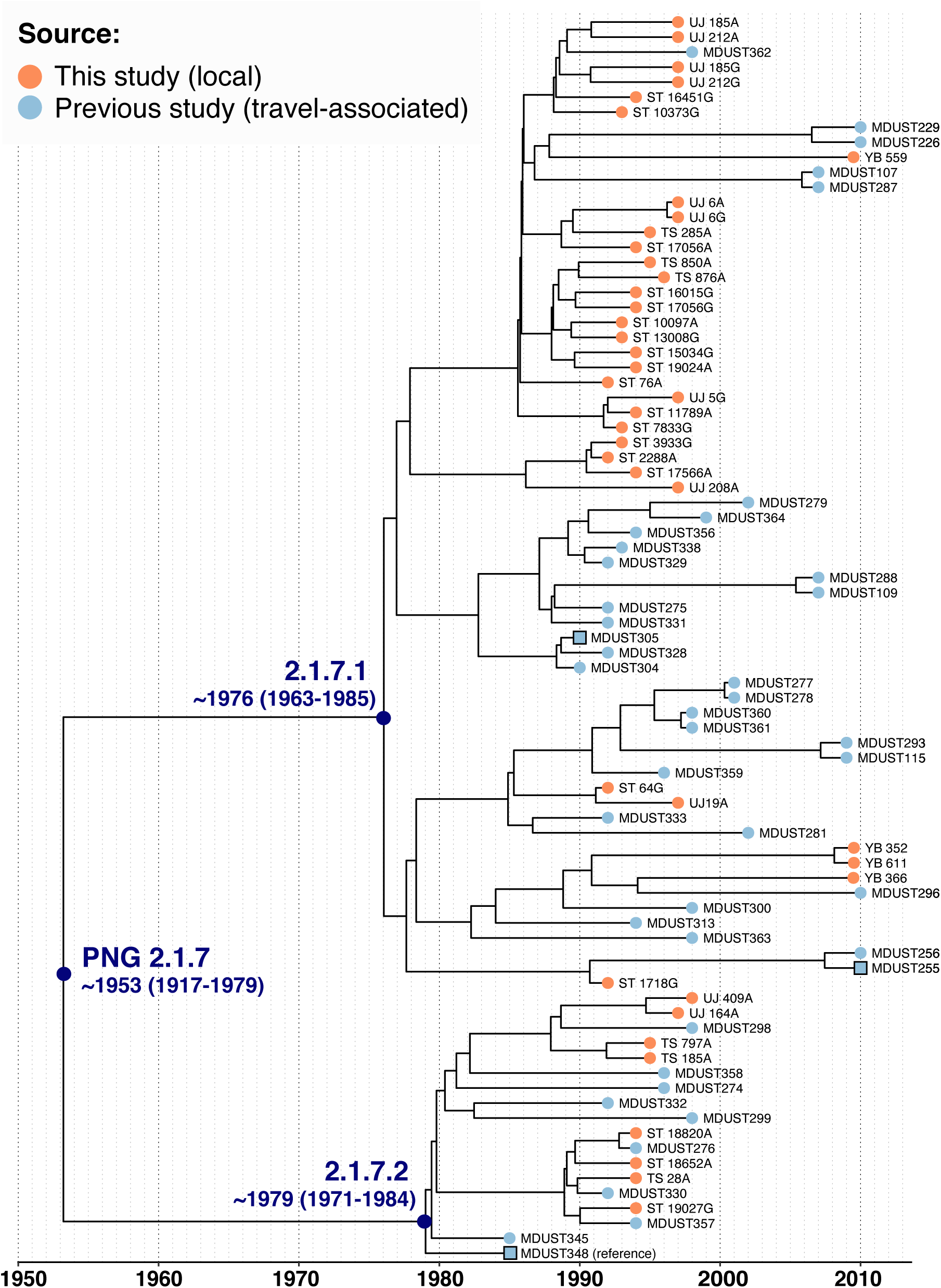
Maximum clade credibility tree inferred from all available (n=83) PNG 2.1.7 *S*. Typhi using BEAST2. Tip colours indicate isolate source as per inset legend, branches are labelled by *S*. Typhi genotype. Square nodes indicate the reference sequence and other completed genomes. Divergence dates for PNG genotype 2.1.7 and sublineages are labelled (95% highest posterior density in brackets). Interactive phylogeny available at: https://microreact.org/project/CYcTnSmjT.

**Fig 2.**
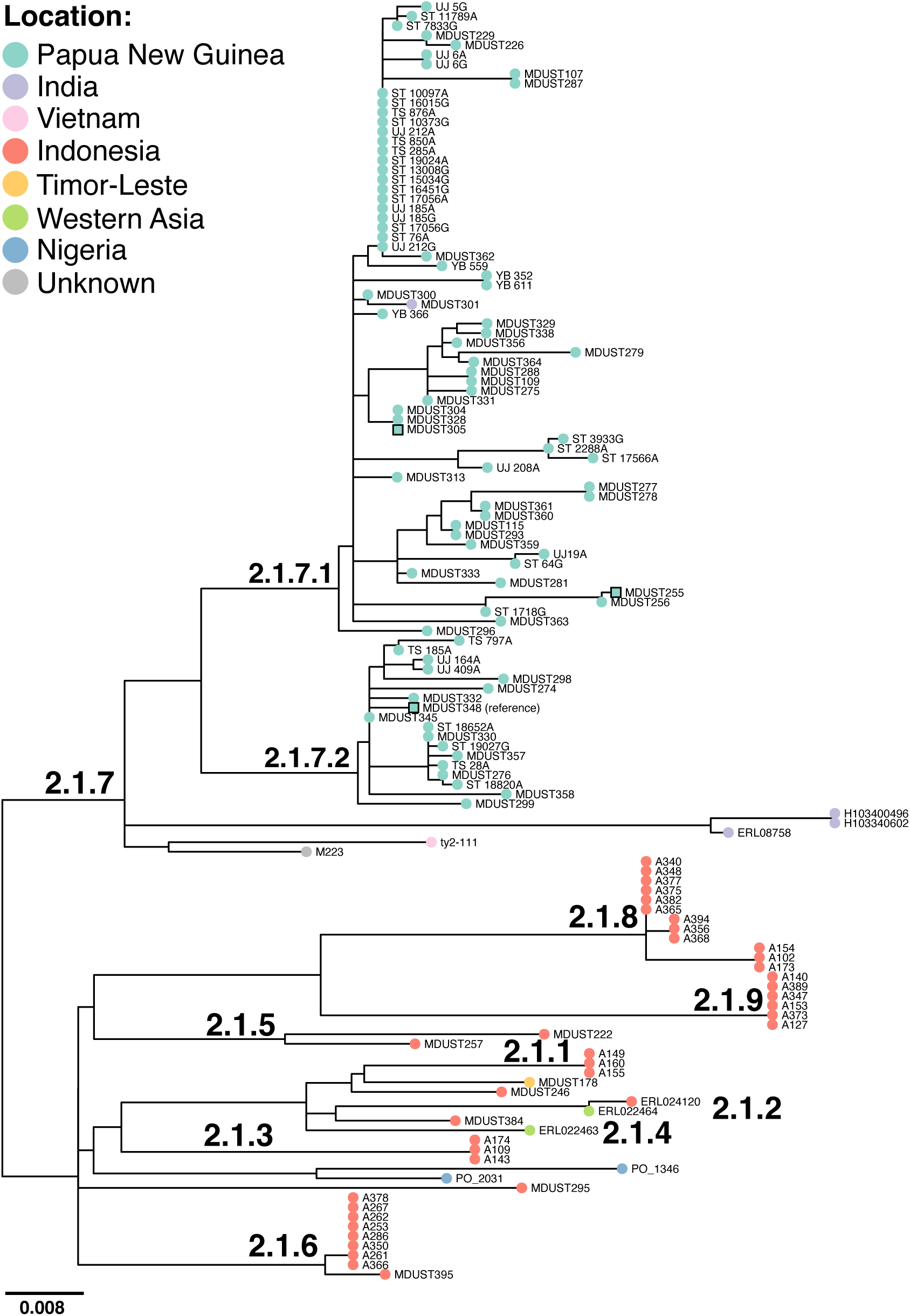
Maximum likelihood phylogeny inferred from all available (n=135) clade 2.1 *S*.Typhi. Tips are coloured by location of origin as per inset legend (for traveller isolates, this is country of travel rather than country of isolation), branches are labelled by *S*. Typhi genotype. Square nodes indicate the position of the reference sequence and other completed genomes. Phylogeny is outgroup rooted with a selection of non-clade 2.1 taxa (see **Methods**). Interactive phylogeny available at: https://microreact.org/project/icGPbF_O8.

Unique SNVs defining PNG genotype 2.1.7 *S*. Typhi sublineages were manually extracted from RedDog allele tables using R. SNVs causing non-synonymous mutations in highly conserved genes were prioritized for sublineage definitions. SNV distances were calculated from the core genome SNV alignment using snp-dists (v0.7.0; available at https://github.com/tseemann/snp-dists).

Assessments of the gene conservation across PNG genotype 2.1.7 *S*. Typhi (**S4 Fig**) were derived from the gene presence/absence matrix output from the RedDog mapping pipeline using the MDUST348 PNG 2.1.7 *S*. Typhi reference genome completed in this study.

### Temporal analysis

To investigate the temporal signal and emergence of genotype 2.1.7 *S*. Typhi in PNG, we used multiple methods. Initially we used TempEst (v1.5.1) [39] to assess the temporal structure of the data (i.e. signals of clock-like behaviour) by conducting a regression analysis of root-to-tip branch distances for the PNG genotype 2.1.7 ML tree inferred using the MDUST348 reference genome **(S2 Fig)** as a function of sampling time. The heuristic residual mean squared method was used for the regression analysis with the best-fitting root option selected. The resultant data from these analyses were visualised in R.

In order to estimate divergence times we analysed the sequence data with BEAST2 [40] (v2.4.7), using the GTR + Γ substitution model and sampling times (tip dates) defined by the year of isolation to calibrate the molecular clock. For n=4 sequences (YB 352, YB 366, YB 559, and YB 611; **S1 Table**), the precise year of isolation was not available, however, as they were known to have been collected between 2009-2010 their date of isolation was estimated with BEAST2 using a tip date sampling prior specifying a uniform distribution between 2009-2010. Models were fit using both constant-coalescent population size and Bayesian skyline tree priors, together with either a strict clock model or a relaxed (uncorrelated lognormal distribution) clock model, to identify the model combination that best fit the data. For all model and tree prior combinations, a chain length of 10,000,000 steps sampling every 5,000 steps was selected[41]. Preliminary analyses highlighted the constant-coalescent population size and relaxed (uncorrelated lognormal) clock models to best fit the data. However, this model combination yielded an implausible divergence date (∼1914, 95% HPD = 1835-1969), compared to those inferred from other model combinations (∼1953, 95% HPD = 1979-1917), and analyses of n=43 genotype 2.1.7 from the previous study alone [27] suggested the Baysian skyline tree prior was better suited to the data. Therefore, the analyses presented here are those using the coalescent Bayesian skyline model together with a relaxed (uncorrelated log normal) clock model. To test the temporal signal we conducted a date-randomisation test whereby sampling times were randomly shuffled, and the analysis re-run 20 times[41,42]. These tests (**S3 Fig**) indicated that the data displayed temporal structure [41].

For the final analysis reported here, a single independent run conducted with a chain length of 10,000,000 states, sampling every 5,000 iterations, was subjected to removal of the first 10% of steps as burn-in with LogCombiner (v2.4.7)[40]. Maximum-clade credibility (MCC) trees were then generated with ‘common ancestor heights’ specified for node heights using TreeAnnotator (v2.4.6)[40]. Effective sample sizes were estimated to be >200 for all parameters reported. All phylogenies were visualised using the R package ggtree (v2.2.4) [43] and the online tool Microreact [44]; an online interactive dated phylogeny is available at https://microreact.org/project/CYcTnSmjT.

### Reference genome assembly and annotation

Hybrid Illumina-Nanopore genome assemblies were generated with Unicycler (v0.4.7) [45] for three PNG genotype 2.1.7 isolates MDUST305, MDUST348, and MDUST255 (see **S2 Fig and S1 Table**). The resultant *de Brujin* assembly graphs were then resolved into single contigs where necessary through manual curation with Bandage (v0.8.1)[46], and annotated with PROKKA (v1.14.0)[47]. Nucleotide pairwise comparisons of the completed genomes were carried out using Mauve [48] (vSnapshot_2015-02-25 1) and visualised using the genoPlotR (v0.8.11) package for R[49].

For analysis of prophage regions detected with PHASTER [37] in hybrid assembled sequences, these were aligned with their closest genetic relative in CT18 (prophage ST35) [50] using Mauve to confirm the prophage region boundaries and insertion site. Prophage regions were then extracted manually from the bacterial chromosome using seqret from the EMBOSS toolkit[51]. Annotations were then transferred to our PNG sequences from the CT18 reference sequence for prophage ST35 using the Rapid Annotation Transfer Tool (RATT)[52], and then manually curated together with PROKKA derived annotations. BLASTn comparisons were then carried out, and visualized using genoPlotR[49].

### AMR detection

The mapping based allele typer SRST2 [53] (v0.1.8) was used to detect the presence of known acquired AMR genes and plasmid replicons using both the ARG-annot [54] and PlasmidFinder [55] databases, respectively. SNVs in the Quinolone Resistance Determining Regions (QRDR) of genes *gyrA, gyrB*, and *parC* that are known to reduce susceptibility to fluoroquinolones in *S*. Typhi [7], and *acrB* mutations associated with azithromycin resistance in typhoidal *Salmonella*[12], were detected using GenoTyphi (**described above**).

### Nucleotide sequence accession numbers

Raw read sequences for the 41 novel isolates from PNG have been deposited in the European Nucleotide Archive under project PRJEB20541; and individual accession numbers are listed in **S1 Table**. Genome assemblies for completed sequences of traveller isolates MDUST348, MDUST255, and MDUST305 were deposited in GenBank under accession numbers TBA, TBA, and TBA respectively (see **S1 Table** for details).

## Results

### Population structure and antimicrobial resistance of S. Typhi in PNG

We combined sequence data from n=41 *S*. Typhi collected locally in PNG and sequenced for this study, with n=45 sequences from a previous study [27] that originate from returned travellers who acquired their infections in PNG. Together these data span 30 years (1980-2010). Travel-associated and locally-collected isolate sequences were intermingled in the inferred phylogeny (**S1 Fig**), and revealed that the population structure of *S*. Typhi during this period was dominated by a single genotype, 2.1.7 (n=84, 98%; isolated 1985 to 2010) which has been present for at least 25 years. Only one other genotype was detected; clade 4.1 (n=2, 2.3%; isolated 1980 and 1986) (**S1 Fig**).

The population structure of genotype 2.1.7 in PNG can be subdivided into two main lineages **(S2 Fig)**, members of which are separated by a median pairwise distance of ∼33 SNVs. We designated these as lineage I (genotype 2.1.7.1) and lineage II (2.1.7.2). In our data, genotype was more common with n=66 (78.6%) members, compared to n=18 (21.4%) 2.1.7.2 members. Interestingly, 2.1.7.2 was not observed after 1998. Genotype 2.1.7.1 can be identified by the presence of a synonymous marker SNV C1503T in gene STY4417 (position 4286788 in CT18), and genotype 2.1.7.2 can be identified by the presence of a synonymous marker SNV C540T in gene STY4106 (position 3967063 in CT18). These genotypes have been added to the GenoTyphi scheme available at http://github.com/katholt/genotyphi[27,35,56].

No known acquired AMR genes, nor mutations associated with either decreased ciprofloxacin susceptibility or azithromycin resistance (see **Methods**) were detected among any of the *S*. Typhi sequences from PNG, indicating this population is sensitive to commonly used antimicrobials. Susceptibility testing data was available for n=40 (88.9%) traveller isolates, all of which tested susceptible to all drugs tested (see **Methods** and **S1 table**).

### Emergence and transmission dynamics of PNG genotype 2.1.7 sublineages

We applied Bayesian phylodynamic analysis to the set of PNG 2.1.7 sequences to estimate the dates of emergence for genotype 2.1.7, and its sublineages 2.1.7.1 and 2.1.7.2 (**Fig 1**, interactive phylogeny available at: https://microreact.org/project/CYcTnSmjT). This analysis yielded a local substitution rate of 0.49 SNVs per genome per year (95% HPD 0.32-0.66) or 1.1×10^−7^ genome-wide substitutions per site per year (95% HPD 7.3×10^−8^-1.5×10^−7^). These data showed temporal structure [41] to support these results (see **Methods** and **S3 Fig**), which were slightly lower than published estimates for *S*. Typhi for genotype 4.3.1 in Nepal (1.7×10^−7^ genome-wide substitutions per site per year, 95% HPD 1.1×10^−7^–2.4×10^−7^)[8], Kenya (1.9×10^−7^, 95% HPD1.5×10^−7^-2.2×10^−7^)[57], and globally (1.42×10^−7^, 95% HPD 1.0×10^−7^ to 1.8×10^−7^)[9]. We estimate that the most recent common ancestor (MRCA) for all PNG genotype 2.1.7 existed circa ∼1953 (95% HPD = 1917-1979) as shown in **Fig 1**. PNG 2.1.7 sublineages appear to have emerged contemporaneously in PNG in the mid-late 1970s, with the MRCA of 2.1.7.1 estimated circa ∼1976 (95% HPD 1963-1985) and the MRCA of 2.1.7.2 around the same time or potentially slightly later, circa ∼1979 (95% HPD 1971-1984).

In order to better understand the relationship of PNG genotype 2.1.7 *S*. Typhi to its nearest relatives in the pathogen population, we inferred a phylogeny of all available genome sequences belonging to its parent clade 2.1 (**Fig 2**, interactive phylogeny available at: https://microreact.org/project/icGPbF_O8). Genotype 2.1.7 isolates were mostly from PNG (n=84, 93.3%). Within genotype 2.1.7, PNG sequences formed a distinct cluster (further subdivided into lineages 2.1.7.1 and 2.1.7.2), with a small number of distant relatives in India (three genotype 2.1.7 isolates separated from PNG isolates by a median pairwise distance of ∼64 SNVs) and Vietnam (one sequence separated by median ∼41 SNVs). The only non-PNG isolate clustering within the PNG sublineages was a single 2.1.7.1 sequence from India (MDUST301 from 1988; **S1 Table and Fig 2**), which was intermingled in the phylogeny with the PNG isolates. This sequence was separated by a distance of ∼3 SNV from its closest PNG relative suggestive of transmission of this genotype from PNG to India on at least one occasion. The other (i.e. non-2.1.7) isolates from clade 2.1 originated almost exclusively from other islands in the Indonesian archipelago (total n=40 isolates, 90.1%; see **Fig 2**), suggesting that the PNG 2.1.7 group emerged from a local pathogen population that had been established in the region for some time.

### Genetic variation among PNG genotype 2.1.7 S. Typhi

As described in **Materials and Methods**, we completed three PNG genotype 2.1.7 genomes to facilitate high resolution analysis of this clone. Three 2.1.7 genomes from PNG *S*. Typhi were selected for completion; two were the oldest sequences from each sublineage available (genotype 2.1.7.1 sequence MDUST305 isolated in 1990, and genotype 2.1.7.2 sequence MDUST348 isolated in 1985). We also completed a recent sequence of the commonest genotype 2.1.7.1 (sequence MDUST255 isolated in 2010). The three completed genomes were markedly similar in size, differing by as little as 4.4 kbp (**Fig 3a**). Further, pairwise nucleotide sequence comparisons revealed high levels of genetic similarity (99-100% coverage, 100% identity), and evidence of genome plasticity with multiple large-scale inversions observed adjacent to repetitive ribosomal RNA (*rrn*) operons (**Fig 3b**).

**Fig 3.**
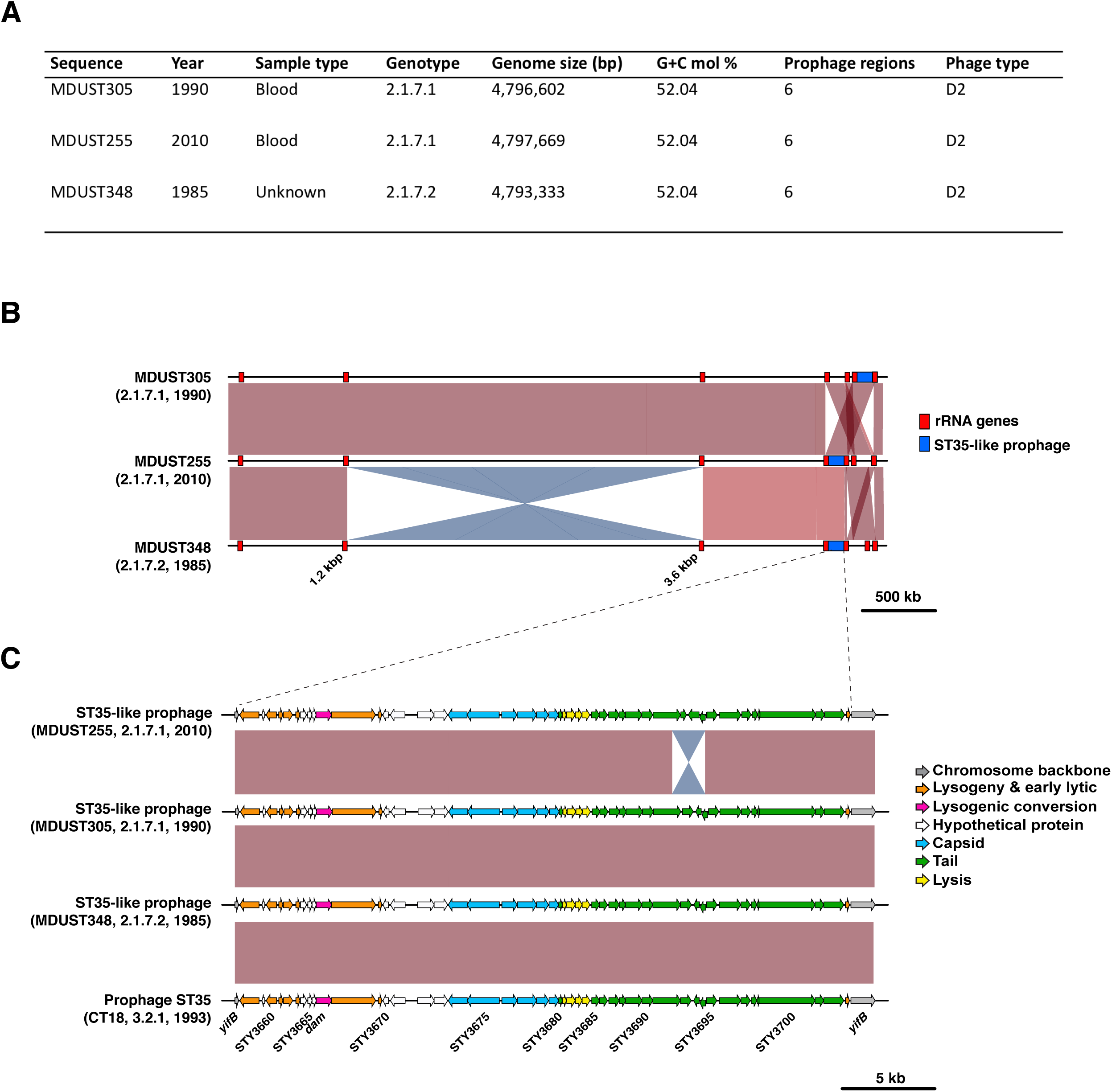
Genome plasticity among PNG *S*. Typhi. **(A) Summary of genetic features of completed PNG 2.1.7 *S*. Typhi genomes. (B) Pairwise nucleotide alignments of PNG 2.1.7 *S*. Typhi**. Shaded regions indicate nucleotide sequence homology, red blocks indicate ribosomal RNA (rRNA) operons, and blue blocks indicate the ST35-like prophage as per the inset legend. **(C) Pairwise alignment of prophage ST35-like elements in PNG *S*. Typhi and the CT18 reference sequence**. Shaded regions indicate nucleotide sequence homology, and genes are coloured by putative function as per the inset legend.

Gene content was highly conserved among both PNG 2.1.7 sublineages (96-100% coverage; **S4 Fig**). Very few deletions were observed, and were not restricted to a particular sublineage. Three sequences, two of genotype 2.1.7.1 (MDUST329 and UJ 212G) and one of 2.1.7. 2 (MDUST332) appeared to have lost the *Salmonella* pathogenicity island 7 (SPI7) locus known to encode genes for Vi polysaccharide capsule formation, type IVB pilli, and bacteriophage SopE. Such deletions are known to occur in the laboratory and did not appear to affect the phage types previously determined for the PNG 2.1.7 traveller isolates as all were of phage type D2. Besides the deletion of prophage SopE within SPI7, no other prophage deletions were apparent.

Examination of detected prophage elements amongst the three completed prophage sequences revealed a smaller ∼1.8 kbp inversion within an ST35-like prophage in the genome sequence of MDUST255 (**Fig 3c**). The inversion was located in the prophage tail gene module carrying homologues of prophage ST35 genes (from reference sequence CT18) encoding for a tail fibre (STY3691), tail fibre assembly chaperone (STY3692), alternate tail fibre tips (STY3693 and STY3694), a site-specific invertase (STY3695), and major tail tube filament structural gene (STY3696)[50]. The inversion, likely mediated by the site-specific invertase (STY3695), appears to have resulted in the formation of an alternative tail fibre protein; a fusion of the N-terminal region of tail fibre protein STY3691 with alternative tail fibre tip protein STY4694. Examination of alignments of read data to reference sequence MDUST348 suggested that this locus was present in both orientations across the PNG 2.1.7 population in line with previous reports[50].

## Discussion

These findings demonstrate that the vast majority of typhoid fever cases in PNG are caused by a single lineage of *S*. Typhi (genotype 2.1.7) which emerged in the Indonesian archipelago in the mid-twentieth century before differentiating into two sublineages in the mid-late 1970s, with genotype 2.1.7.2 no longer appearing common after 1998. These findings correlate well with the broad epidemiology of typhoid fever in PNG. Typhoid fever was sporadically reported in PNG in the 1960s, becoming more commonly diagnosed in the 1970s and then becoming endemic in the PNG highlands in the 1980s [16,58]. Thus, the divergence dates for PNG *S*. Typhi 2.1.7 sublineages correspond to the approximate time when typhoid fever became endemic. *S*. Typhi genotype 2.1.7 appears to have persisted within PNG since this time, and was dominant among both returning travellers and locally detected cases over the course of 30 years of genomic surveillance. We observed little evidence of genetic variation among the genotype 2.1.7 population, with infrequent gene deletions (**S4 Fig**), no obvious evidence of prophage or plasmid acquisitions (**Fig 3** and **S4 Fig**), and a lower substitution rate compared to the widespread H58 (4.3.1) genotypes associated with intercontinental transmission and AMR[8,9,57]. Notably, our data suggest that AMR has not emerged among the *S*. Typhi population in this setting, with both genotype and phenotype data revealing all isolates to be sensitive to former first line drugs (chloramphenicol, ampicillin and co-trimoxazole), as well as fluoroquinolones, third generation cephalosporins, and macrolides.

The AMR sensitivity of the PNG 2.1.7 genotype and the long term stability of this population is in sharp contrast to many other endemic settings, e.g. South Asia where infections are commonly driven by H58 genotypes[9], both MDR and reduced susceptibility to fluoroquinolones are common [7,8,59,60], and resistance to third generation cephalosporins[10,11], and azithromycin [12–15,61] have also emerged.

We observed limited evidence of inter-country transmission, with no obvious recent introductions into PNG from outside. A small number of genotype 2.1.7 isolates were found outside PNG in India and Vietnam, but these were mostly distantly related to the PNG lineages and we observed only one possible case of transmission of PNG genotype 2.1.7 *S*. Typhi into India (travel-associated case of lineage 2.1.7.1 isolated in 1988 **Fig 2**). It is possible, however, that this individual may also have visited PNG as well as India in their travels with only the later recorded. The limited international movement of *S*. Typhi in and out of PNG may partially explain why AMR has not been observed in this setting, in the form of the arrival of H58 or other drug resistant genotypes of *S*. Typhi.

Among PNG 2.1.7 populations, the lack of emergence of AMR may be influenced by a lack of local selective pressures related to antimicrobial usage. Tracking antimicrobial usage is challenging in PNG, but evidence of sub-optimal administration of antibiotics has been reported[62]. Approximately 73% of outpatients with non-malarial febrile illness are administered antibiotics, commonly contrary to the relevant prescription guidelines.

Amoxicillin, trimethoprim-sulfamethoxazole and benzylpenicillin collectively accounted for ∼75% of antibiotic administrations, demonstrating the high dependence on β-lactams in PNG, but little exposure to cephalosporins. Data pertaining to other Gram-negative pathogens in PNG are limited. A recent investigation of *Shigella* from PNG and the Pacific region revealed this pathogen to have resistance to ampicillin, chloramphenicol, tetracycline, and trimethoprim-sulfamethoxazole, with resistance more common in bacteria isolated since 2010. Resistance to cephalosporins (ceftriaxone) and ciprofloxacin was not detected in *Shigella* from PNG[63]. On the basis of observations in *Shigella*, resistance to first-line antibiotics could emerge in other gastrointestinal pathogens in PNG, but exposure, and therefore resistance to, cephalosporins and second-generation quinolones (e.g. ciprofloxacin) is unlikely.

Genome variation reported previously in the PNG *S*. Typhi population [18] appears to be primarily mediated by large-scale re-arrangements adjacent to repetitive *rrn* operons (**Fig 3b**), rather than a diverse range of genotypes or the acquisition of mobile genetic elements such as prophages or plasmids consistent with previous reports[18,19]. Rearrangements between the *rrn* operons have been described previously in *S*. Typhi and are driven by homologous recombination [64,65]. This has previously been postulated to cause sequence variation (e.g. diverse ribotypes) among PNG *S*. Typhi[19,20], and our data support this hypothesis. Recent long-read data suggests that rearrangements around the *rrn* operons are relatively common in *S*. Typhi[66], and appear to be associated with host persistence and carriage in the gallbladder[67]. We also observed inversions in the tail morphogenesis gene module of the ST35-like prophage. These inversions are also well described and are likely a mechanism for generating variation in tail fibre genes that are known to be involved with the attachment stage of viral replication[68]. Such variation is known to cause alteration of phage host ranges[50,69].

This study is not without limitations, primarily relating to the challenges associated with obtaining clinical isolates of *S*. Typhi. There is no ongoing routine surveillance conducted in PNG, nor is there routine blood culture for diagnosis of febrile illness. Consequently, locally collected isolates are obtained primarily through targeted studies. Most isolates analysed in this study originated from Eastern Highlands Province, where the PNG Institute of Medical Research is headquartered and is thus the site of most studies. Eastern Highlands Province is one of 22 provinces in the country (albeit the second most populated province), thus there is a lack of geographical distribution of isolates. Given most isolates originate from targeted studies, the temporal distribution of isolates is intermittent. Moreover, the lack of ongoing surveillance precluded contemporary isolates (from the past 10 year) being collected and thus included in this study. However, as locally collected and travel associated cases both demonstrated a lack of AMR and similar genotype frequencies, in line with limited previously published sequences[21], these combined data appear representative of major trends among the *S*. Typhi population in this setting.

Overall, our data highlight that unlike other recently examined settings in Kenya, Zimbabwe and other regions of Sub-Saharan Africa, as well as South Asian settings in India, Nepal, Bangladesh and Pakistan [8,10,57,59,60,70–73] the *S*. Typhi populations in PNG appear sensitive to a diverse range of antimicrobials with former first line drugs, fluoroquinolones, third generation cephalosporins and macrolides all remaining viable treatment options prior to the introduction of Vi-conjugate vaccines and improvements to water, sanitation and hygiene (WaSH) related infrastructure. However, despite this, further ongoing surveillance is required to monitor for the potential emergence or introduction of AMR *S*. Typhi in this setting.

## Data Availability

Genomic data are submitted to the European Nucleotide Archive (ENA) and accession numbers are available in the supplementary tables.

## Acknowledgements

The authors wish to thank Sebastian Duchene for useful discussions regarding phylodynamic analyses.

## Supporting Information Captions

**S1 Fig. Maximum likelihood phylogeny for all available *S*. Typhi originating in PNG**. Tip colours indicate the source of the sequence as per the inset legend. Branches are labelled by *S*. Typhi genotype. Square nodes indicate the position of the reference sequence and other completed genomes.

**S2 Fig. Maximum likelihood phylogeny for all available PNG genotype 2.1.7 *S*. Typhi**. Tip colours indicate the source of the sequence as per the inset legend, and branches are labelled by genotype. Square nodes indicate the position of the reference sequence and other completed genomes. A Bayesian dated tree inferred using the same alignment and year of isolation is shown in **Fig 1**.

**S3 Fig. Temporal Analysis of PNG genotype 2.1.7 *S*. Typhi. (A) Tempest regression of root-to-tip distances as a function of time**, with the root of the tree selected using heuristic residual mean squared. Each point represents a tip in the maximum likelihood phylogenetic tree shown in **S2 Fig**. The slope is a crude estimate of the annual substitution rate for the SNV alignment, the x-intercept corresponds to the age of the root node, and R^2^ is a measure of clock-like behaviour among the data. **(B) Date randomisation test results** with the right most box plot showing the posterior substitution rate estimate from the SNV alignment of the data with the correct sampling times, and the remaining 20 boxplots showing the posterior distributions of the rate estimate from replicate runs where the dates were subjected to randomisation. The data are considered to have strong temporal structure if the estimates using the correct dates do not overlap with those where the dates were randomised.

**S4 Fig. Genetic conservation among PNG 2.1.7 *S*. Typhi**. Branches are labelled by genotype. Coloured square nodes indicate the position of the reference sequence and other completed genomes. Heatmap shows the presence or absence of all annotated genes in the reference sequence as per the inset legend. Features of interest are highlighted with red boxes and labels.

**S1 Table. Accession numbers and data for *S*. Typhi sequences used in this study (Excel file)**

**S2 Table. Excluded repeat and phage regions in PNG MDUST348 2.1.7.2 completed reference sequence (Excel file)**

**S3 Table. Outgroups used for phylogenetic tree rooting (Excel file)**

