## Supplementary figures and images for "Whole genome sequence analysis of *Salmonella* Typhi in Papua New Guinea reveals an established population of genotype 2.1.7 sensitive to antimicrobials"

### S1 Fig

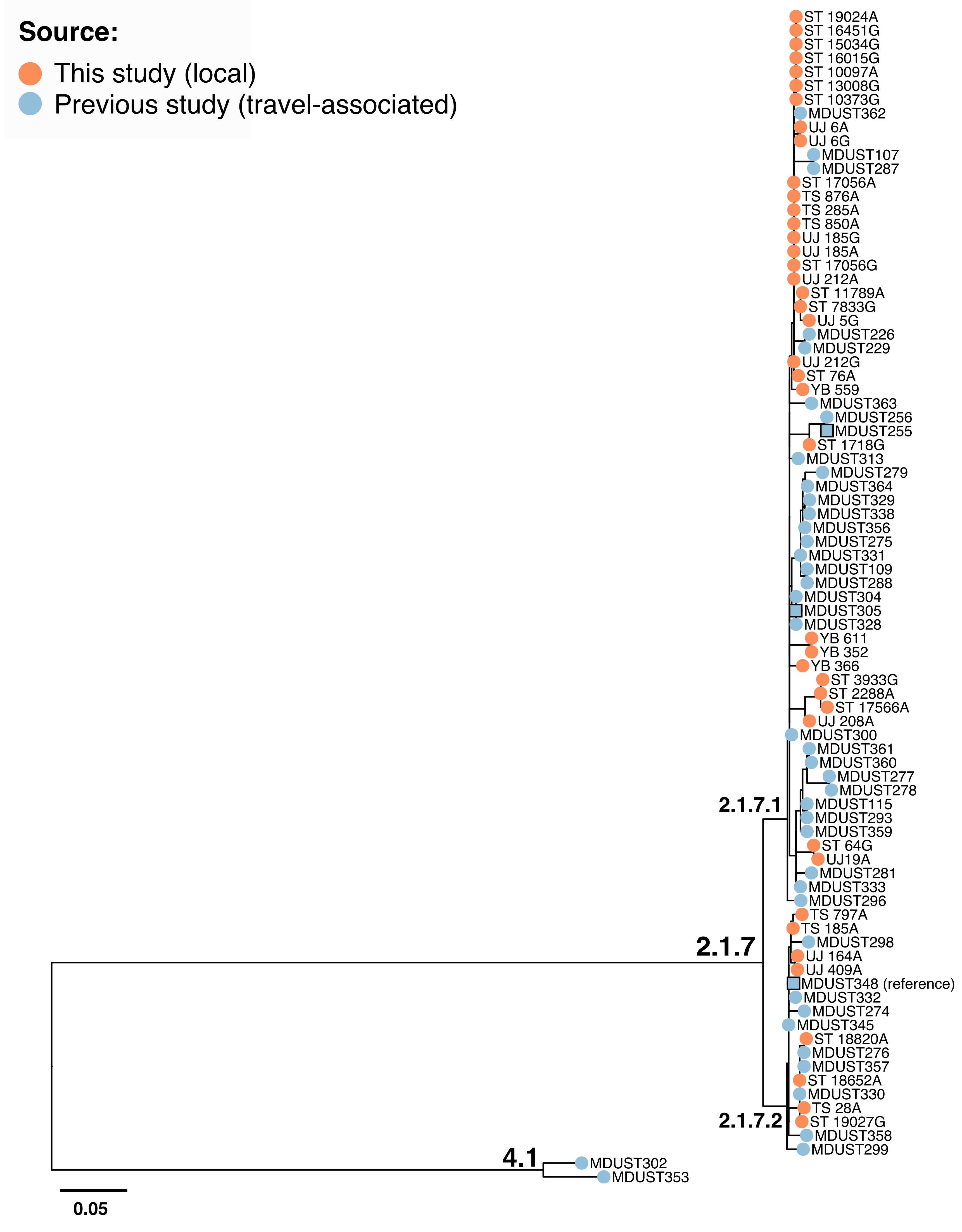

### S2 Fig

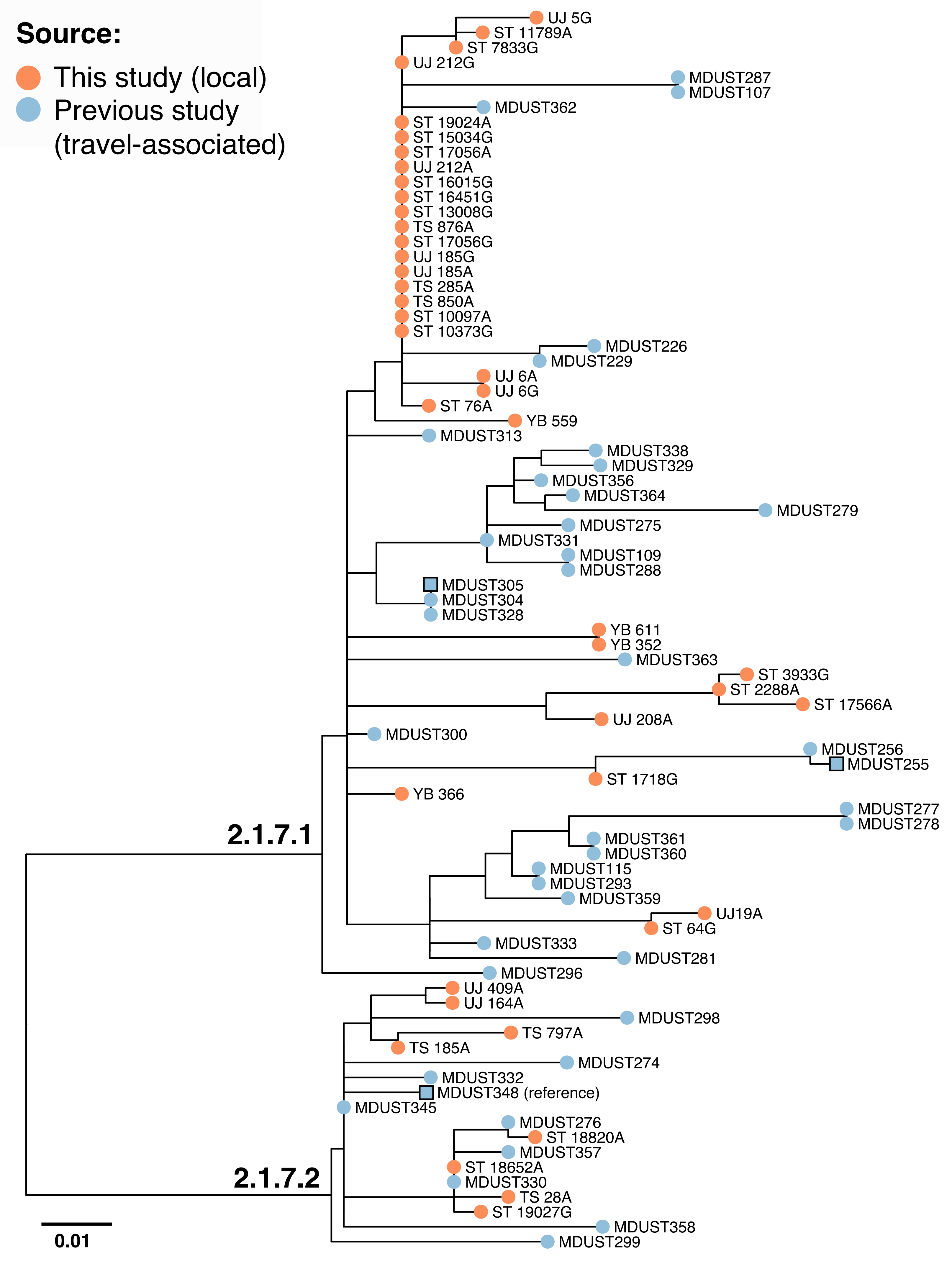

### S3 Fig

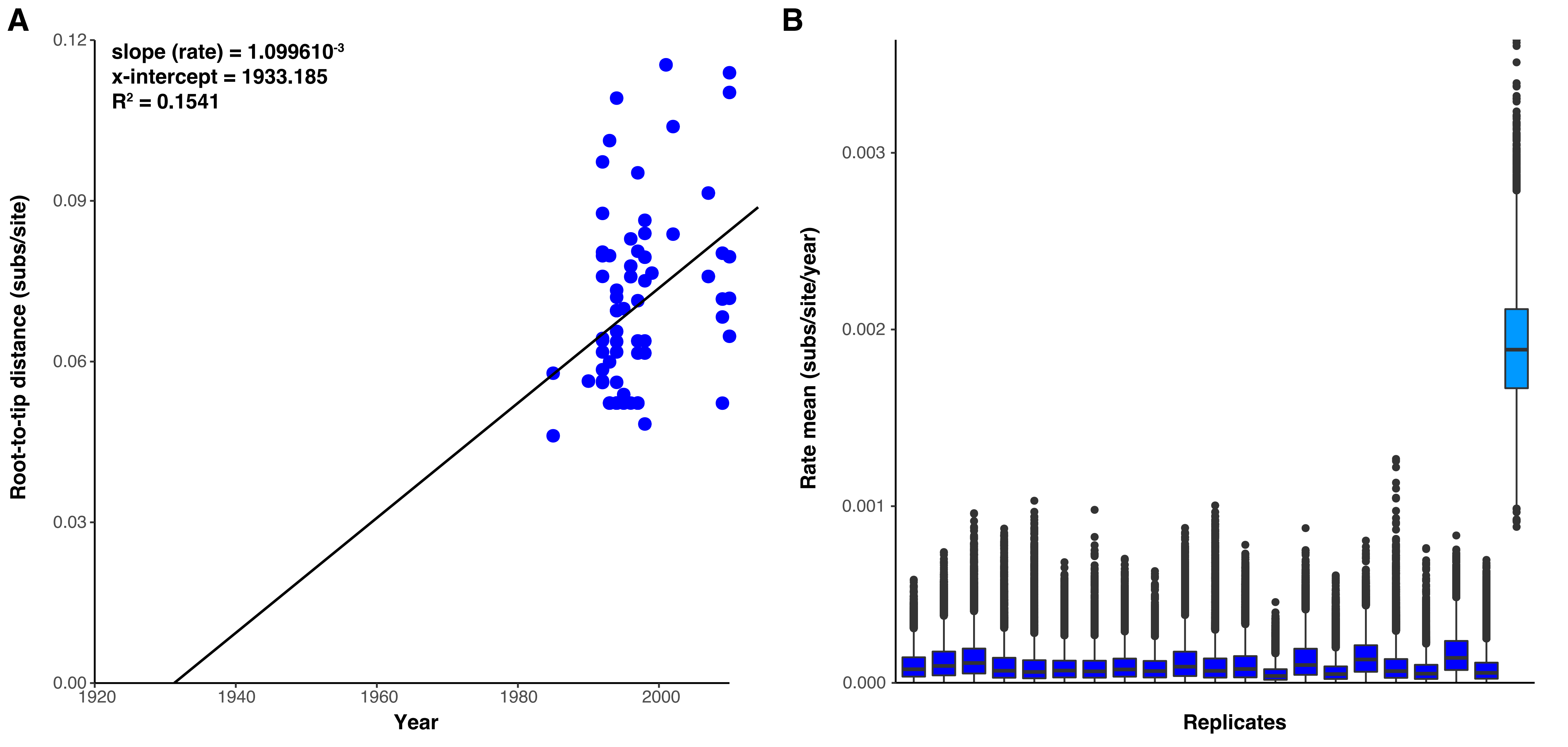

### S4 Fig

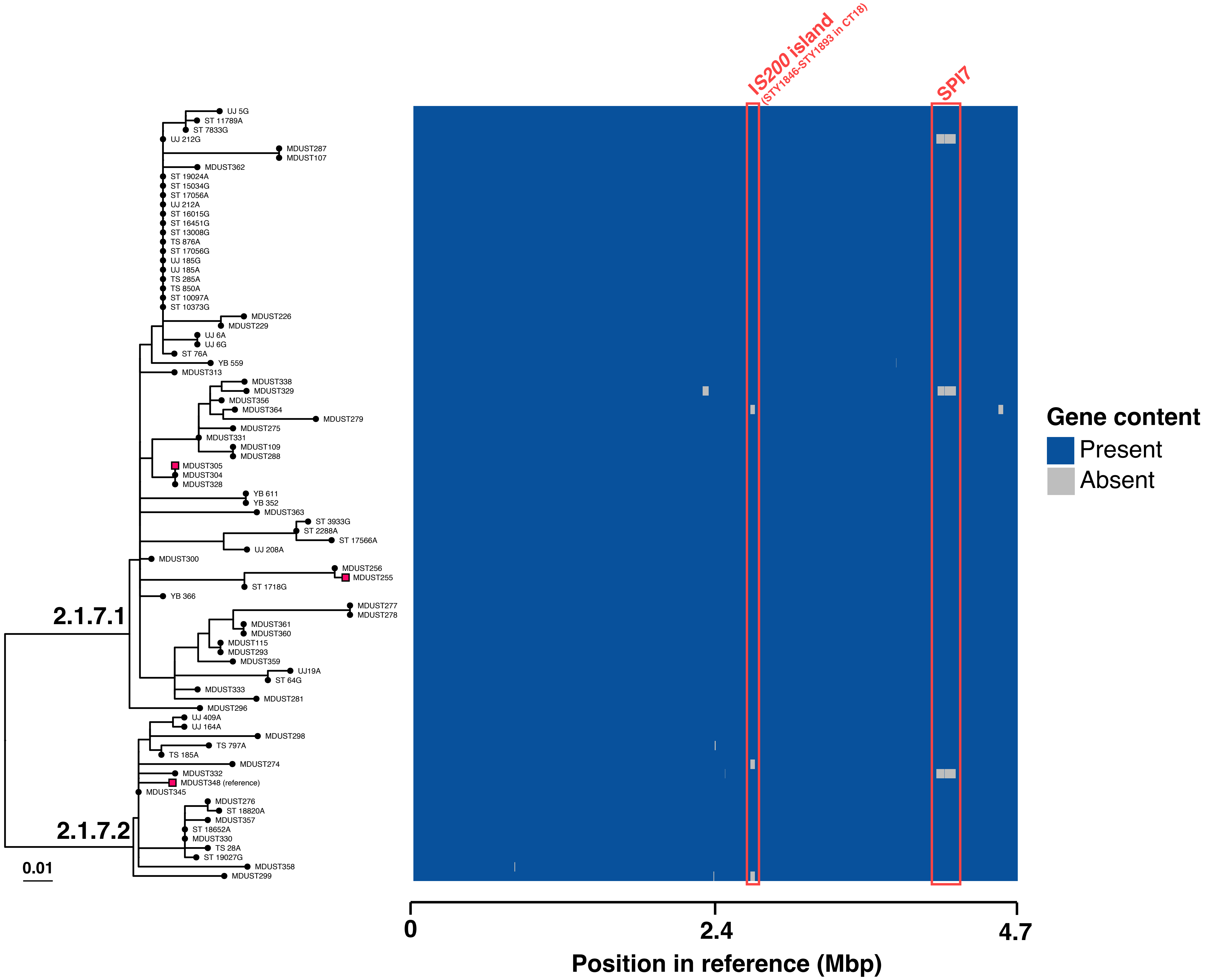
